# Pan-disease blood protein profiles of rheumatic autoimmune diseases

**DOI:** 10.64898/2026.02.05.26345462

**Authors:** Josefin Kenrick, Charlotta Preger, María Bueno Álvez, Alejandra Ulloa, Göran Bergström, Antonella Notarnicola, Begum Horuluoglu, Anna Smed-Sörensen, Anna Färnert, Anna Norrby-Teglund, Iva Gunnarsson, Marie Wahren-Herlenius, Marie Holmqvist, Leonid Padyukov, Karine Chemin, Lina Marcela Diaz-Gallo, Ingrid E. Lundberg, Elisabet Svenungsson, Vivianne Malmström, Lars Klareskog, Sofia Bergström, Mathias Uhlén, Peter Nilsson, Fredrik Edfors, Elisa Pin

**Author notes:** Corresponding author: **Correspondence to**: PhD Elisa Pin, SciLifeLab, Div of Affinity Proteomics, Dept of Protein Science, School of Engineering Sciences in Chemistry, Biotechnology and Health (CBH), KTH Royal Institute of Technology. These authors contributed equally to this work and share senior authorship (Fredrik Edfors, Elisa Pin).

## Abstract

Systemic autoimmune rheumatic diseases (SARDs) are a heterogeneous group of autoimmune conditions characterized by immune system dysregulation leading to chronic inflammation and tissue damage. The overlapping clinical manifestations make differential diagnosis challenging, highlighting the need for novel biomarkers to facilitate early diagnosis, stratification, and personalized treatment. Five SARDs including idiopathic inflammatory myopathies (n=210), rheumatoid arthritis (n=84), systemic sclerosis (n=100), Sjögren disease (n=99), and systemic lupus erythematosus (n=99), as well as healthy controls (n=400) and controls with acute infectious diseases (n=218) were selected for plasma protein profiling using Olink Explore 1536. Proteins with known association to SARDs as well as novel associations were identified through differential abundance analysis and machine learning. This explorative cross-sectional study demonstrates the importance of a pan-disease approach to biomarker identification within and between the five SARDs. NPX boxplots from this study are available open-access through the Human Protein Atlas, facilitating further plasma-proteome research on autoimmune diseases.

## Introduction

Systemic autoimmune rheumatic diseases (SARDs) is an umbrella term for several heterogeneous rheumatic disorders, all characterized by an immune system imbalance, which implies systemic organ and tissue involvement with overlapping clinical manifestations and serological features^1^. Despite advancements, diagnosis of SARDs can be complicated and requires a multifaceted approach involving clinical evaluation from specialists alongside laboratory tests. SARDs include among others rheumatoid arthritis (RA), systemic lupus erythematosus (SLE), systemic sclerosis (SSc), Sjögren disease (SjD), and idiopathic inflammatory myopathies (IIM). Among these conditions, challenges in differential diagnosis stem from the wide range of clinical manifestations, involvement of multiple organs, and overlapping symptoms and signs such as fatigue, joint pain, and inflammation. The therapeutic opportunities are rapidly increasing for most of these diseases, with growing potential choice among different therapies for the various rheumatic diseases and their different subsets. There is thus a high need for additional biomarkers that can distinguish between these diseases and their subsets and contribute to the development of personalized, targeted therapies.

While autoantibodies are the hallmark of many SARDs and useful for diagnosis, they do not always reflect the ongoing inflammatory processes. Further, most currently used inflammatory markers are far too unspecific to contribute to the differential diagnosis. Genetics remains useful for understanding disease pathogenesis but is so far not useful in clinical diagnostics. Blood-based protein biomarkers provide a valuable complement to currently available tools for diagnosis and evaluation of disease activity, and they also offer insights into the often-unclear biological processes underlying the different diseases. New blood-based protein biomarkers may contribute both to molecularly driven differential diagnoses and to the identification of potential therapeutic targets.

Affinity proteomics has already been successfully applied to identify candidate biomarker panels for systemic lupus erythematosus (SLE), systemic sclerosis (SSc)^2,3,4^, and rheumatoid arthritis (RA)^5^. However, most of these studies focus on single diseases and comparisons with healthy controls, therefore not providing information on the overall disease specificity and usefulness for differential diagnosis of the candidate biomarkers. To ensure their reliability and clinical utility, plasma biomarkers should be evaluated in a pan-disease setting across diverse patient populations and pathological conditions to assess their specificity^6^. Within the SARDs context, a recent study screened the level of 161 mainly immunoregulatory proteins in the blood of patients with SLE, ANCA-associated systemic vasculitis, RA, and Sjögren disease (SjD) using antibody arrays, and reported a small number (n<10) of disease-specific proteins that could differentiate the diseases^7^.

In the present study, we applied Proximity Extension Assay (PEA) to screen 1472 proteins in the plasma of 592 patients with SARDs as part of the Human Disease Blood Atlas – namely SLE, RA, SjD, SSc, and IIM – as well as 400 healthy controls and 218 patient samples from three acute infectious diseases. This study provides a comprehensive analysis of the plasma proteome of five SARDs, identifies single proteins and protein panels that can differentiate between different SARDs and from controls. Protein levels are available from this study in the open access Human Protein Atlas (www.proteinatlas.org) as a resource for further research.

## Methods

### Samples and patients

The study included 592 patients with systemic autoimmune rheumatic diseases (SARDs) enrolled at the Rheumatology clinic of Karolinska University Hospital in Stockholm, Sweden and 400 healthy controls between ages 50 to 65 as part of the Swedish CArdioPulmonary bioImage Study (SCAPIS) study^8^ (Fig. 1a). To serve as an additional set of controls, 218 samples from patients with streptococcal soft tissue infection (n=77), acute influenza virus infection (n=98), and malaria (n=43) were included, with the aim to account for non-autoimmune inflammation markers. The SARDs included in this study were IIM (n=210), RA (n=84), SjD (n=99), SSc (n=100), and SLE (n=99) (Fig. 1a). Plasma EDTA was collected in K2 EDTA tubes, prepared through centrifugation at 2.400 x g for seven minutes, and subsequently stored at – 80°C. Most samples were collected at time of diagnosis and pre-treatment. Classification criteria for the diseases was based on the EULAR/ACR 2017 criteria for IIM^9^, the EULAR/ACR 1987^10^ or 2010^11^ criteria for RA, the EULAR/ACR 2016 criteria for SjD^12^, the EULAR/ACR 2013^14^ criteria for SSc, and the ACR 1982 revised classification and the 2012 SLICC criteria for SLE^12,14^.

**Fig. 1:**
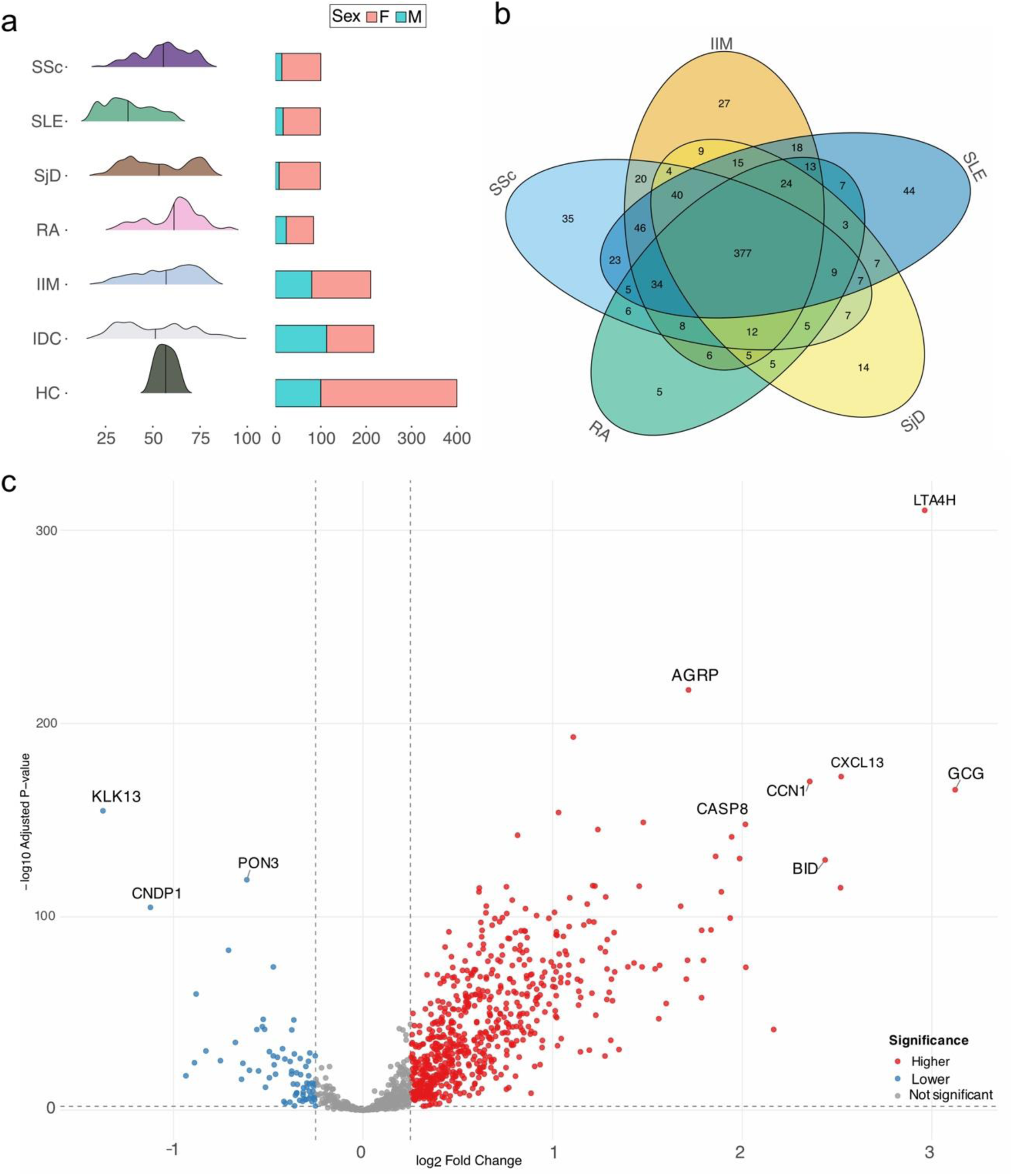
Cohort characteristics and autoimmune protein signatures. **a**) Age and sex distribution for systemic sclerosis (SSc), systemic lupus erythematosus (SLE), Sjögren disease (SjD), rheumatoid arthritis (RA), idiopathic inflammatory myopathies (IIM), infectious disease control (IDC), and healthy control (HC). **b**) Venn diagram showing the number of overlapping proteins from each individual SARD that had higher levels (adjusted p-value < 0.01, logFC > 0.25) compared to the healthy control. **c**) Volcano plot highlighting proteins with differential levels in autoimmune cohort in comparison to healthy controls. Proteins with significantly higher levels (adjusted p-value < 0.01, logFC > 0.25) in autoimmune cohort are shown in red, and lower levels (adjusted p-value < 0.01, logFC < –0.25) against healthy in blue.

Study participants were enrolled and samples collected according to the ethical principles outlined in the Declaration of Helsinki. Information regarding ethical permits and hospital affiliations can be found in the supplementary data (Supp data 1).

### Proximity Extension Assay

Protein levels were measured in plasma using the Olink Explore 1536 platform as part of the Human Disease Blood Atlas program^15^. Samples were fully randomized and normalized following standard plate protocol defined by Olink Proteomics AB (Uppsala, Sweden), as described in Álvez et al^6^. Protein levels are shown as normalized protein expression (NPX) units on a log2 scale. Eight proteins were identified by quality control (QC) at Olink Proteomics AB as having a high-dose hook effect, and those were removed alongside 304 proteins that had a strong correlation (rho > 0.7) with them. The protein PCOLCE was excluded due to a high number of assay QC warnings. Samples were removed if over 50% of NPX values had QC warnings. When needed, k-nearest neighbor imputation was used for Uniform Manifold Approximation and Projection (UMAP) and machine learning.

### Statistical Rationale

Differential abundance analysis was performed using *limma* (v3.62.2) package in R (v4.4.1), correcting for both age and sex^16^. Benjamini-Hochberg multiple hypothesis correction was used^17^. Each autoimmune disease was compared to a grouping of the other four autoimmune diseases, which served as a SARD control group. Each single SARD was also compared to both healthy controls and controls with infectious diseases following the same differential abundance procedure. Adjusted p-values as well as log2 of the fold change (logFC) for each protein and comparison was calculated. Due to the large number of differentially abundant proteins within the comparisons, an adjusted p-value cut-off of 0.01 was chosen.

Machine learning classification was performed in a supervised manner using multinomial GLMnet lasso with *tidymodels* (v1.3.0) package. NPX values were normalized, near zero variance filter was applied, and missing values were imputed. Nested cross-validation was used across five independent folds, split into training and testing sets for only SARD samples. Each training set contained an inner loop of ten validation and training sets. Validation sets were used for tuning hyperparameters. Receiver operating characteristic (ROC) curves were computed for each fold, and the average area under the curve (AUC) was taken across the five folds. A confusion matrix was calculated based on classification of the test set from each fold. The training set was used to determine importance of individual features on the model using raw model coefficients and the *vip* (v0.4.1) package.

Gene set enrichment analysis was performed using *clusterProfiler* (v4.16.0) package in R. All graphs were made using *ggplot2* (v3.5.1) in R. Uniform Manifold Approximation and Projection (UMAP) was made using *uwot* (v0.2.3) package in R^18^.

## Results

### Protein differential abundance analysis: SARDs vs healthy controls and infectious disease controls

Initially, we analyzed all five SARDs versus healthy controls and infectious disease controls to identify plasma proteins that are unique to SARDs. In total, 377 proteins showed increased levels (adjusted p-value < 0.01, logFC > 0.25) in every autoimmune disease against healthy controls, with further overlap in the inter-SARD comparisons (Fig. 1b, Supplementary Fig. S1). Fewer proteins were found to have lower levels (adjusted p-value < 0.01, logFC < – 0.25) than the healthy controls, with 16 proteins identified as shared across all SARDs (Supplementary Fig. S2).

In the overarching comparison of the grouped SARDs versus healthy controls, proteins involved in inflammatory response and response to oxygen containing compounds had higher levels based on gene enrichment analysis (Supplementary Fig. S3). Among proteins that had higher levels in the grouped autoimmune diseases compared to healthy controls we detected proteins involved in inflammatory and immune pathways including leukotriene A4 hydrolase (LTA4H), cellular communication network factor 1 (CCN1), and C-X-C motif chemokine ligand 13 (CXCL13) (Fig. 1c).

When comparing the grouped autoimmune diseases with the infectious disease controls, gene set enrichment analysis revealed that the SARD group had higher levels of proteins involved in cell adhesion and morphogenesis, among other functions (Supplementary Fig. S4a-b). Among single proteins, cytokines and inflammatory proteins such as colony stimulating factor 3 (CSF3) and interleukin 10 (IL-10) had lower levels in the autoimmune group in comparison to the infectious disease controls (Supplementary Fig. S4c).

### Protein differential abundance analysis: each SARD vs other SARDs

The next stage of the analysis included differential analysis of each systemic autoimmune rheumatic disease against the other four SARDs. Idiopathic inflammatory myopathies (IIM) was the disease with the highest absolute number of proteins with increased levels against all others, with rheumatoid arthritis (RA) having the highest absolute number of proteins with decreased levels (Fig. 2a-b, Table 1). Several proteins showed to be increased in level in more than one disease against other grouped SARDs, with the largest overlap between systemic sclerosis (SSc) and IIM (76 proteins). (Fig. 2b).

**Fig. 2:**
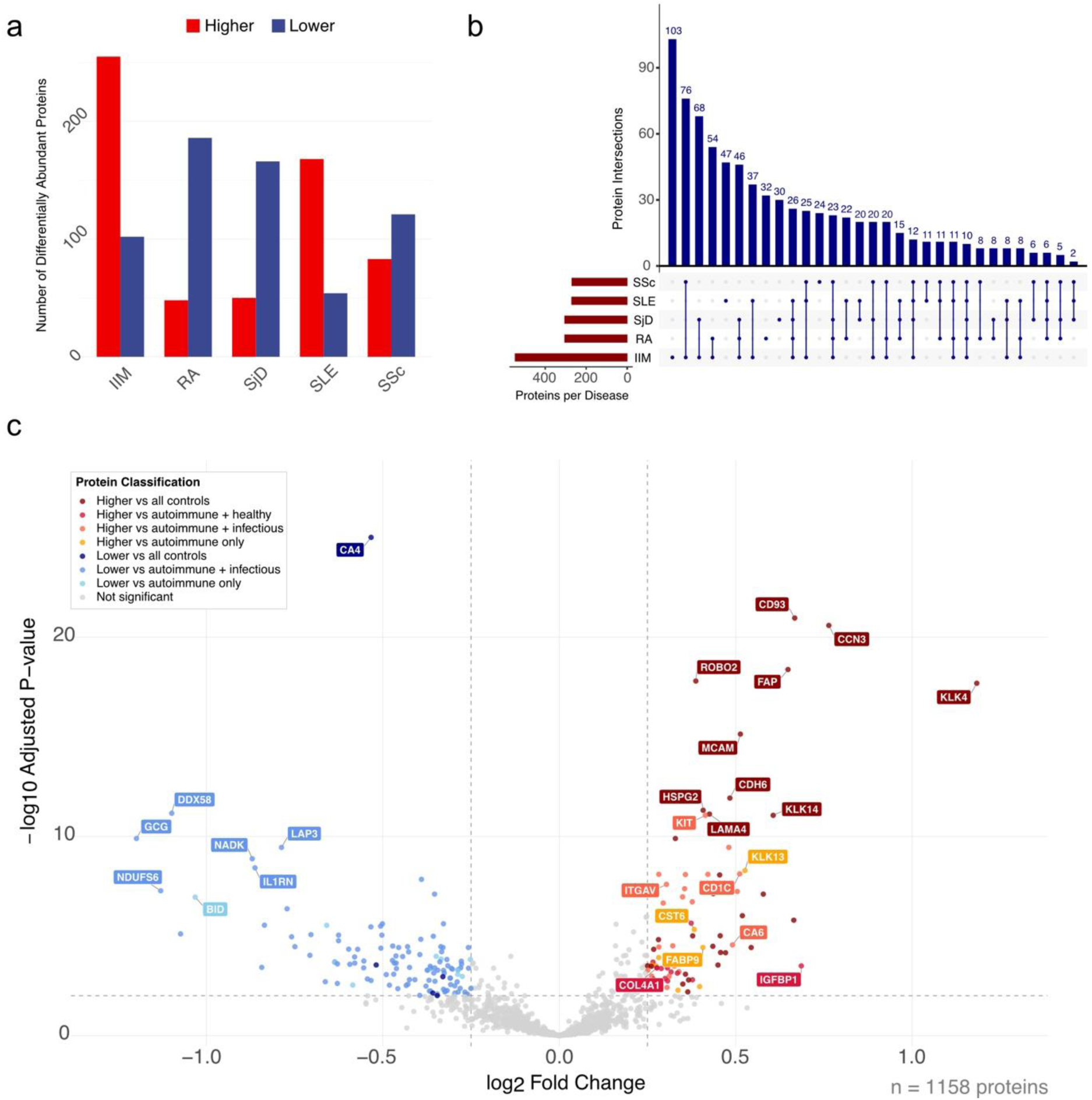
Differentially abundant plasma proteins across diseases. **a**) Bar plot showing the number of proteins identified by differential analysis for each SARD using adjusted p-value cutoff of 0.01, logFC > or < 0, red = higher levels, blue = lower levels. **b**) Upset plot showing proteins with higher abundance in each SARD based on cutoff adjusted p-value < 0.01, logFC > 0.25. **c**) Volcano plot for comparison of systemic sclerosis to the grouped autoimmune diseases, annotated by comparisons to healthy and grouped infectious diseases.

**Table 1:** Number of proteins significantly increased or decreased in single-SARD compared with other grouped SARDs (adjusted p-value < 0.01)

| Disease | Increased levels | Decreased levels |
| --- | --- | --- |
| IIM | 266 | 101 |
| RA | 23 | 138 |
| SjD | 33 | 134 |
| SLE | 135 | 44 |
| SSc | 85 | 127 |

To identify proteins specific for each single SARD, each elevated protein was annotated based on the result of the comparison against healthy and infectious disease controls (Fig. 2c, Supplementary Fig. S5-8). An example of a protein that may be disease specific in SSc is kallikrein-4 (KLK4), which was elevated in comparison to the other SARDs as well as both control groups. In contrast, the protein mast/stem cell growth factor receptor, KIT, was elevated in SSc compared to the other SARDs and the grouped infectious diseases but showed no significant difference relative to healthy controls. Finally, insulin-like growth factor binding protein 1 (IGFBP1) was higher in SSc than in other SARDs and healthy controls, but not compared to the infectious disease controls, suggesting it may be related to non-specific autoimmune inflammatory features (Fig. 2c). Similar patterns were found for all five SARDs (Supplementary Fig. S5-S8). The results here will focus on proteins that were elevated in a specific autoimmune disease compared to all other SARDs.

In systemic lupus erythematosus (SLE) we identified higher abundance for immune related proteins, such as microfibril associated protein 5 (MFAP5) and proprotein convertase subtilisin/kexin type 9 (PCSK9) (Fig. 3, Supplementary Fig. S5). In addition, TNFSF11 (also known as RANK ligand), was elevated in SLE compared to all other groups (Fig. 3). Among the proteins with increased abundance, SLE shared 11 with SSc (e.g. RBP2, FABP2) and 37 with IIM (e.g. VSIG4, IL-15).

**Fig. 3:**
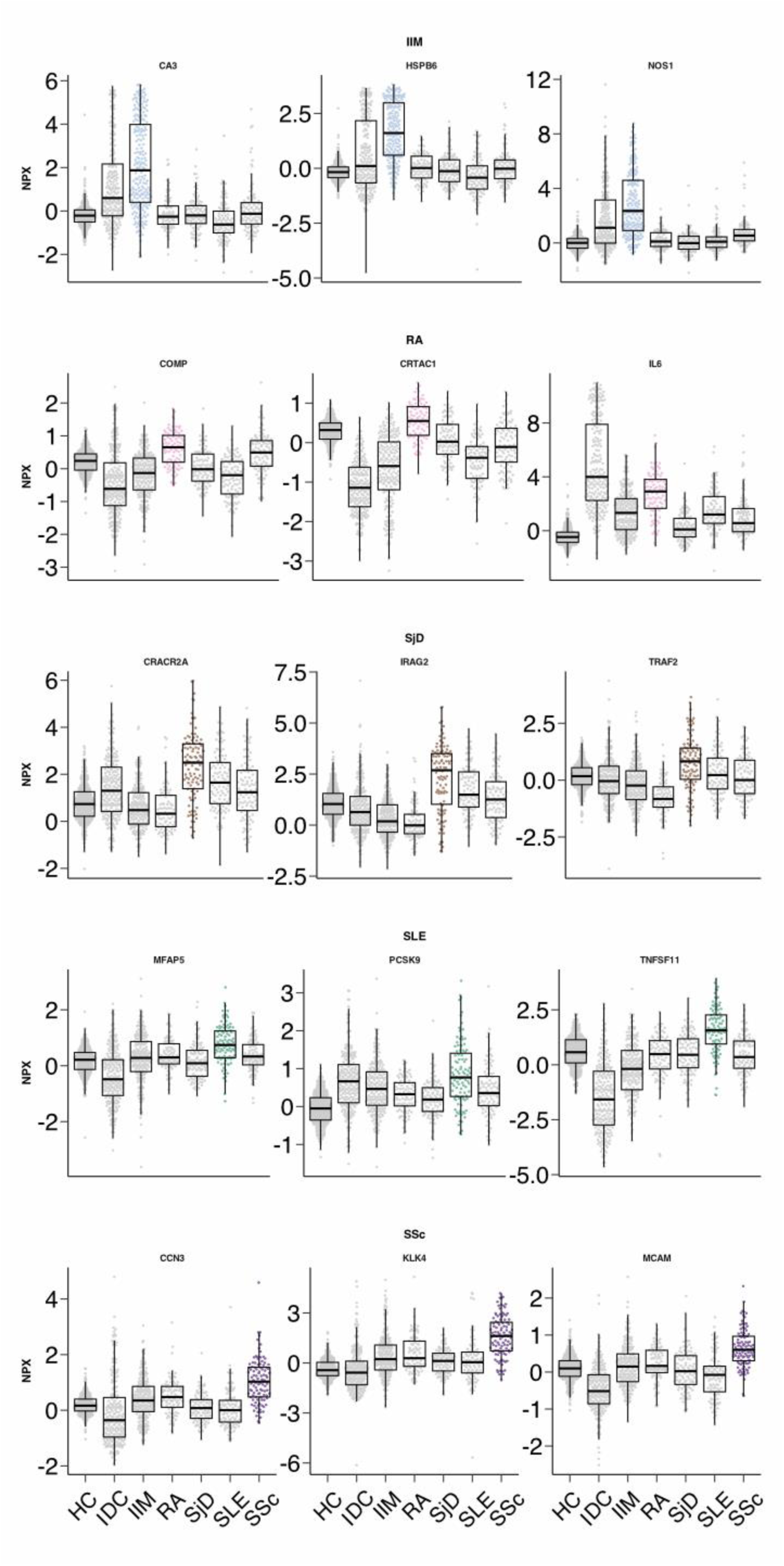
Plasma protein profiles across autoimmune diseases and controls. Each row represents one disease with box plots showcasing a disease-specific pattern, having higher protein levels (normalized protein expression, NPX) than the other SARDs as well as both controls. Each dot represents one individual. IIM, idiopathic inflammatory myopathies in blue, RA, rheumatoid arthritis in pink, SjD, Sjögren disease in brown, SLE, systemic lupus erythematous in green and SSc, systemic sclerosis in purple.

Differential analysis for RA identified proteins involved in deep zone cartilage, such as cartilage acidic protein 1 (CRTAC1), and cartilage oligomeric matrix protein (COMP), despite having fewer differentially abundant proteins compared to the other SARDs (Fig. 3). Additionally, the established marker and therapeutic target interleukin 6 (IL6) was identified, despite being unsurprisingly higher in the infectious diseases^19^ (Fig. 3, Supplementary Fig. S4c, Supplementary Fig. S6).

In Sjögren disease (SjD), the transporter protein inositol 1,4,5-triphosphate receptor associated 2 (IRAG2) had higher levels in comparison to the other SARDs. It has a connection to the immune system through its suggested role in the major histocompatibility complex (MHC), indicating a potential link to autoimmunity^20^ (Fig. 3, Supplementary Fig. S7). Other proteins found elevated in SjD were tumor necrosis factor ligand superfamily member 11 (TRAF2), a TNFR receptor, and EF-hand calcium-binding domain-containing protein 4B (CRACR2A) a calcium binding protein.

As stated, IIM had numerous differentially abundant proteins, and notably many of those with higher levels were specific to muscle tissue, such as nitric oxide synthase 1 (NOS1), carbonic anhydrase 3 (CA3), and heat shock protein family B (small) member 6 (HSPB6)^21^. (Fig. 3, Supplementary Fig. S8).

Previously mentioned proteins KLK4 and MCAM, found elevated in SSc, have roles in enamel formation and cell adhesion, respectively^22,23^. Lastly, cellular communication network factor 3 (CCN3), which impacts a variety of tissue-building functions including cell proliferation, adhesion, migration, and differentiation, was found to be elevated in SSc compared to all controls (Fig. 3)^24^.

### Machine learning classification

Machine learning multiclassification was applied with a nested cross-validation strategy, a supervised approach using a GLMnet lasso regression model to classify all SARDs simultaneously. This helped identify complex patterns between the different diseases, and allowed for selection and ranking of protein features, thus complementing the differential abundance analysis.

Across the five independent folds, ROC curves indicated high performance of the model, with average AUCs on the test set ranging from 0.98 – 0.99 (Fig. 4a). From the confusion matrix, the disease most often misclassified was SjD, still with only a few cases (Fig. 4b). From the training set, we identified the top features for classification of each disease (Fig. 4c). These top features, or important proteins, are identified via the absolute value of raw model coefficients ranging from 0 to 1.1643. Notably, protein leukotriene A4 hydrolase (LTA4H) was considered important for the classification of both IIM and SjD, while inositol 1,4,5-triphosphate receptor associated 2 (IRAG2) appeared for SjD, SLE, and RA. These results confirm the higher level of IRAG2 found in SjD and SLE compared to the healthy and infectious disease controls in the differential abundance analysis (Fig. 2c). Similarly, LTA4H was identified in the differential abundance as having highest levels when compared to HC for all SARDs. The performance of the classifier was high, with mostly predicted cases matching with true cases. From the GLMnet results, the proteins included in the classifier derived especially from SjD (138), followed by IIM, SLE, SSc, and RA (Fig. 4c, Supp. Data). Using the top 10 proteins deemed important for the classification of each disease, a total of 47 proteins were selected; the UMAP generated using the dataset of 47 proteins showed improved separation of disease clusters compared to using the whole dataset as input (Fig. 4d-e).

**Fig. 4.**
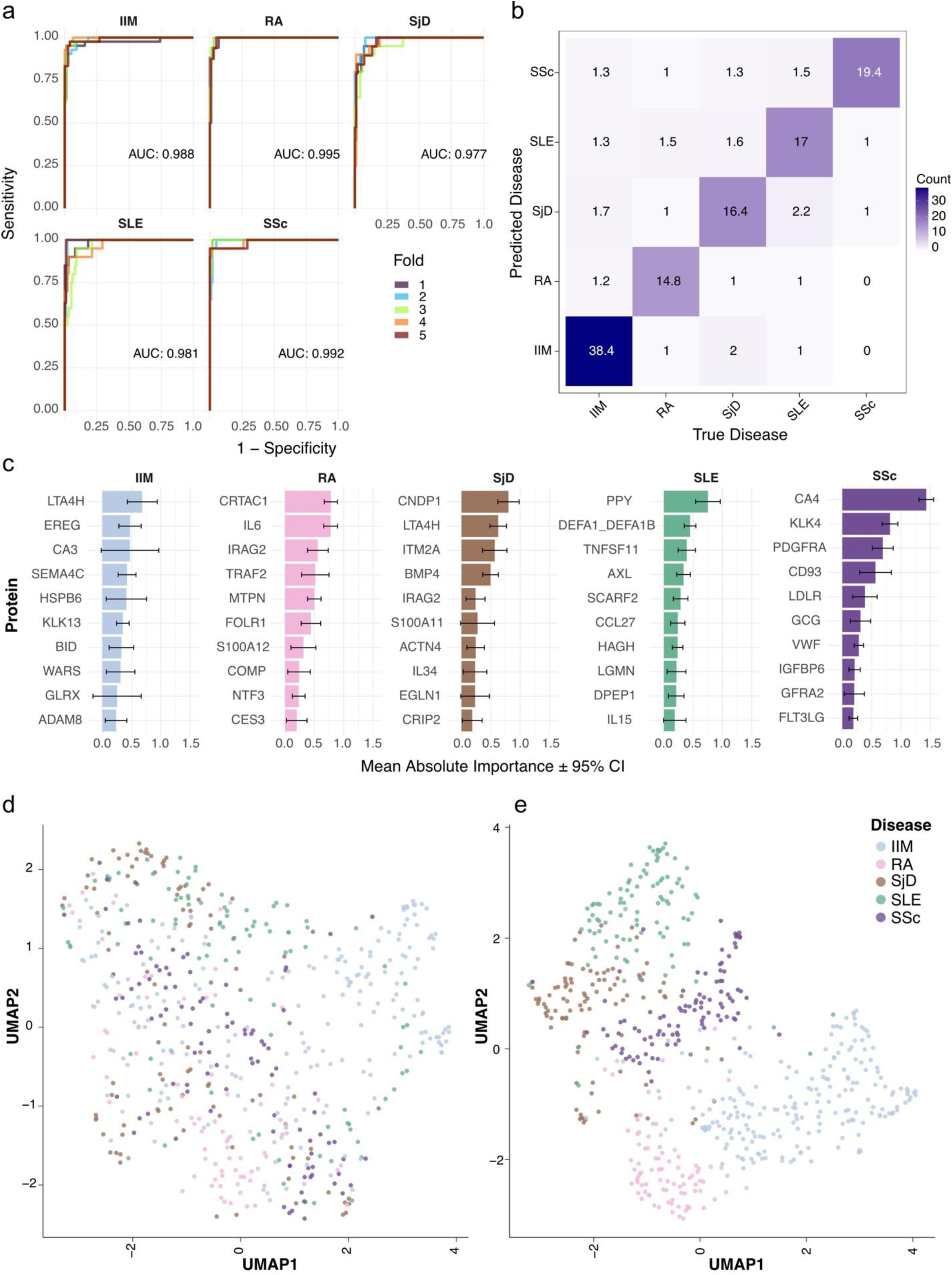
Pan-autoimmune classification using machine learning. **a**) ROC curves of model performance across the five folds with the average AUC for each disease. **b**) Confusion matrix across the folds with the average count of misclassified cases. **c**) Mean absolute importance with 95% confidence interval of the top 10 important proteins by disease from the five folds. **d**) UMAP dimensionality reduction plots shown with all proteins in the dataset and, **e**) after filtering for the top 10 features deemed important for each disease by machine learning (n = 47).

### Combining differential abundance and machine learning for disease-specific protein profiles

A comparison of the results from the differential abundance and machine learning analysis was performed with the aim to select top ranked proteins showing increased levels in one specific SARD compared to other SARDs and both healthy and infectious disease controls, therefore showing the highest level of specificity. Results showed that IIM had the highest overlap of proteins identified by both differential analysis and machine learning, followed by SLE and then SSc, RA, and SjD. Twelve proteins were identified as having higher levels than all controls based on differential abundance analysis and identified via machine learning in all five independent folds (Table 2). This corresponds to two to four proteins per SARD, with IRAG2 being selected in both SjD and SLE.

**Table 2:** Most relevant proteins elevated per disease in comparison to SARDs and consistently selected as important by machine learning.

| Disease | Gene name | Protein name | Uniprot ID | Disease vs Healthy | Disease vs Infectious disease | Machine learning mean importance |
| --- | --- | --- | --- | --- | --- | --- |
|  |  |  |  | logFC (adjusted p-value) | logFC (adjusted p-value) |  |
| IIM | LTA4H | Leukotriene A-4 hydrolase | P09960 | 3.74 (3e-26) | 0.89 (3e-11) | 0.64 ± 0.38 |
|  | CA3<sup>25</sup> | Carbonic anhydrase 3 | P07451 | 2.4 (3e-48) | 0.77 (5e-05) | 0.48 ± 0.41 |
|  | EREG | Proepiregulin | O14944 | 1.82 (9e-53) | 0.60 (7e-05) | 0.47 ± 0.31 |
| RA | CRTAC1<sup>26</sup> | Cartilage acidic protein 1 | Q9NQ79 | 0.20 (2e-3) | 1.39 (1e-26) | 0.82 ± 0.21 |
|  | COMP<sup>27</sup> | Cartilage oligomeric matrix protein | P49747 | 0.37 (1e-07) | 0.77 (4e-10) | 0.23 ± 0.17 |
| SjD | IRAG2* | Inositol 1,4,5-triphosphate receptor associated 2 | Q12912 | 1.44 (7e-12) | 1.48 (1e-11) | 0.27 ± 0.17 |
| SLE | AXL<sup>28,29,30,31</sup> | Tyrosine-protein kinase receptor UFO | P30530 | 0.75 (2e-21) | 0.37 (8e-06) | 0.36 ± 0.16 |
|  | TNFSF11 | Tumor necrosis factor ligand superfamily member 11 | O14788 | 0.66 (2e-4) | 2.65 (2e-29) | 0.33 ± 0.18 |
|  | IRAG2* | Inositol 1,4,5-triphosphate receptor associated 2 | Q12912 | 1.18 (4e-09) | 0.66 (2e-3) | 0.29 ± 0.12 |
| SSc | KLK4<sup>32</sup> | Kallikrein-4 | Q9Y5K2 | 2 (7e-34) | 1.84 (7e-22) | 0.89 ± 0.06 |
|  | CD93 | Complement component C1q receptor | Q9NPY3 | 1.12 (1e-30) | 0.98 (6e-20) | 0.58 ± 0.28 |
|  | FABP2 | Fatty acid-binding protein, intestinal | P12104 | 0.35 (5e-3) | 1.3 (2e-10) | 0.15±0.12 |

*\*This protein was identified as important in more than one disease.*

Altogether, our results provide novel insight into the plasma proteome of five SARDs, showing many proteins with different levels across autoimmune disorders, and with IIM and SLE having the highest number of proteins elevated in plasma when compared to other SARDs and controls. We demonstrate the relevance of using infectious disease controls in addition to healthy controls, particularly in the context of autoimmunity, when strong inflammatory responses might share characteristics with an acute infection. We showed that using the top proteins identified by machine learning allowed us to distinguish the SARDs from each other. Lastly, we present a short list of proteins selected both by differential abundance analysis and machine learning as the most specific proteins for each single disease.

## Discussion

This study leveraged proximity extension assay (PEA) to analyze the plasma proteome in a large cohort of patients with five different SARDs in a pan-autoimmune disease setting. The results of differential abundance and machine learning analyses demonstrated that despite having clinical and proteomic similarities, we were able to identify a subgroup of proteins specific for each of the SARDs.

There were many differentially abundant proteins, some with increased abundance and some with decreased in each disease compared to all other SARDs. In this study, we focused primarily on those with increased levels in each of the studied diseases respectively, as these would be more easily translated into diagnostic biomarkers.

Systemic lupus erythematosus (SLE) and idiopathic inflammatory myopathies (IIM) showed the greatest number of proteins with elevated levels in the differential abundance analysis. In SLE, dysregulation of apoptosis and an insufficient clearance of intracellular proteins and nuclear debris have been reported to be involved in the pathogenesis of the disease^33^. Thus, this could be an explanation for the higher levels of proteins found in the plasma proteome of SLE patients. Regarding IIM, muscle fiber necrosis and regeneration is characteristic of most patients with inflammatory active disease, which could result in the release of more proteins into the plasma.

Several members of the matrix metalloproteinase (MMP) family were identified in our study, with increased levels in SLE. The association between MMP and SLE has been suggested before^33,34^. In addition, the MMP related protein MFAP5, which aids in fibroblast function and scar formation, was also identified in association with SLE through both methods^35^. Neutrophil defensin 1 (DEFA1_DEFA1B), an anti-microbial peptide found in the epithelia of mucosal surfaces, and tyrosine-protein kinase receptor UFO (AXL) both have known roles in SLE and these were among the top proteins for SLE in machine learning and differential abundance^36,37,38,39,40^. Interestingly, both tumor necrosis factor receptor superfamily member 11A (TNFRSF11A aka RANK), and its ligand TNFSF11, part of the NF-k-beta pathway and critical for bone density, were found to have significantly higher levels in SLE compared to the other autoimmune diseases^41^.

Several proteins in IIM had higher levels against the infectious disease controls (IDC), most notably those enriched in skeletal muscle tissue^21^. These proteins ranged from having roles in muscle function (HSPB6) to cholesterol synthesis (IDI2)^42,43^. Other key proteins identified include potential autoantigens, such as tryptophanyl-tRNA synthetase (TrpRS/WARS1). TrpRS is a member of the aminoacyl-tRNA synthetases, a family with several known autoantigens in IIM^44^. In addition, NADH:ubiquinone oxidoreductase subunit S6 (NDUFS6), a mitochondrial protein connected to complex I in the respiratory chain, was one of the proteins that showed a higher level in IIM in our study^45^. This is of interest in connection to a recent study from our group which found autoantibodies against NDUFA11, another protein part of the same complex^46^.

In contrast, many of the higher abundance proteins in IIM appeared in comparison to the other SARDs but had lower levels than the infectious disease controls, such as guanylate binding protein 2 (GBP2), involved in innate immunity, and chemokine ligands CXCL10 and CCL7^47^. These proteins involved in immunity and inflammation are higher in IIM than other SARDs, possibly suggesting higher systemic inflammatory levels than other autoimmune diseases.

Several of the proteins with higher levels in systemic sclerosis (SSc) are involved in tissue synthesis, such as KLK4 of the kallikrein family and CCN3 involved in fibrosis. Other common categories included proteins expressed in endothelial cells, such as MCAM/CD146 and CD93^48,49,50,51^. The soluble version of CD93 has been shown to correlate with disease severity and activity of SSc, which we identified via both differential analysis and machine learning methods^52^. Fibroblast activation protein (FAP) showed high levels in SSc in our study, likely reflecting the involvement of fibroblasts in mechanisms of fibrosis and tissue repair underlying SSc^53^. However, FAP is better known by its role as a tumor marker^54^. The interplay between SSc and cancer is well established^55^ and evident in the results from identification of multiple cancer-related proteins including FAP as well as CDH6^56^, CD93^57^, CCN3^58^, MCAM^23^, KLK4^59^.

For Sjögren disease (SjD), many of the proteins identified were novel in relation to the disease but already reported to be of relevance in cancer biology. Specifically, syntaxin 16 (STX16^60^), TNF receptor associated factor 2 (TRAF2^61^), and calcium release activated channel regulator 2A (CRACR2A^62^) have all been implicated in lymphoid cell and immune related processes and were identified through both differential analysis and machine learning.

The machine learning generated high AUCs and low level of misclassification between the diseases. Additionally, using the top ranked proteins identified from the machine learning, we were able to visibly improve the separation between the different diseases in a high dimensional space, suggesting relevance of these proteins in differentiating between the SARDs.

Our study demonstrated the importance of using a pan-disease approach and inclusion of healthy and infectious disease controls to examine the proteome of SARDs in order to better understand the biology underlying these complex and oftentimes co-occurring diseases and identifying disease-specific proteins as candidate markers. Furthermore, the combination of differential analysis and machine learning is a powerful technique to identify robust candidate markers.

The broad scope and ambitious aim of this study naturally lend to limitations. Firstly, the choice of the control group is crucial for identifying targets, and in this vein, it is important to validate our data using independent healthy and disease cohorts. Furthermore, the number of patients in each cohort is imbalanced, and therefore machine learning analysis should be interpreted with caution. We utilized hyperparameter tuning and cross validation to accommodate for this, but there are still risks of overfitting.

Notably, all the rheumatic diseases investigated in this study can be divided into subsets that are different concerning clinical as well as molecular features. Future studies warrant analysis to address differences between serological and clinical subgroups, as well as disease status based on activity.

Despite the high sensitivity of the proximity extension assays, the broad dynamic range poses a challenge in the accurate measurement of proteins with extreme high or low concentration in blood.

In the context of autoimmune diseases, there are often autoantibodies in the blood. These may interfere with the immunoassay by binding to the protein target of the autoantibody itself or binding to the antibody used for detection, such as in the case of rheumatoid factor^63^. This is worth investigating in a future study.

While in this study we focused on proteins with increased abundance in specific diseases, this decision could be limiting as other proteins may hold valuable insights into the underlying biology of these autoimmune diseases.

As stated, these diseases are overlapping and share many clinical manifestations and are sometimes co-occurring. To get a better overall understanding of the disease mechanisms and pathogenesis and to move towards disease re-classification and more tailored treatment, it will be of interest in a future study to use a combination of clinical manifestations, autoantibody patterns, and protein biomarkers to identify novel molecularly defined disease subsets for future precision medicine efforts.

## Conclusions

Overall, our analysis confirmed the high similarity between the different systemic autoimmune rheumatic diseases on the protein level, which is not surprising given the frequent clinical overlap of some manifestations but also the common co-occurrence of some of these SARDs. Despite this, we identified individual plasma proteins that showed disease specificity, as well as protein combinations enabling discrimination among all five diseases. These protein profiles might aid to distinguish unique biological mechanisms of these diseases. We demonstrated that a pan-disease perspective is critical for identifying SARD-specific biomarkers that are not solely due to inflammation. This data, also available open access through the Human Protein Atlas, represent a resource for further studies. In the future, elucidating differences between subtypes or clinical manifestations of disease is vital for further understanding underlying biology of these SARDs.

## Acknowledgments

We thank the entire staff of the Human Protein Atlas program and the Science for Life Laboratory (SciLifeLab) for their valuable contributions and SciLifeLab Affinity Proteomics at Uppsala University and the National Genomic Infrastructure Uppsala (NGI), supported by SciLifeLab, the Swedish Research Council and the Knut and Alice Wallenberg Foundation, for aiding in protein analyses. We also thank Per Eriksson and Klev Diamanti for their support in analyzing the Olink data. We are grateful to the patients who contributed with blood samples, enabling this research. We acknowledge the funding sources that supported the cohorts and facilitated sample collection, as well as everyone who contributed to patient enrolment and sample collection. This work was supported by the WCPR grant from Knut and Alice Wallenberg Foundation and the SciLifeLab & Wallenberg Data Driven Life Science Program (grant: KAW 2020.0239).

## Ethics Declaration

The research described in this study has been performed in accordance with the Declaration of Helsinki. Ethical approval was obtained from the appropriate regional ethics review boards in Stockholm, Gothenburg, and Lund (Sweden) for the myositis, rheumatoid arthritis, Sjögren’s disease, systemic sclerosis, and healthy control cohorts, and from ethics committees in Copenhagen (Denmark), Vest (Norway), Stockholm, and Gothenburg (Sweden) for the infectious disease control cohorts. Detailed information with protocol numbers are included in Supplementary table 1. All participants received detailed study information and provided written informed consent prior to inclusion in the study.

## Author contributions

Conceptualization and study design: JK, CP, MU, PN, FE, EP. Data analysis: JK, MBA, AU. Data analysis support: CP, EP. Resources: GB, AN, BH, ASS, AF, AN-T, IG, MW-H, MH, LP, KC, LMDG, IEL, ES, VM, LK, SB, MU, PN, FE, EP. Writing – Original Draft: JK, CP, EP. Writing – Review & Editing: MBA, AU, GB, AN, BH, ASS, AF, AN-T, IG, MW-H, MH, LP, KC, LMDG, IEL, ES, VM, LK, SB, MU, PN, FE. Visualization: JK, CP, EP. Supervision: MU, PN, FE, EP. Project administration: LK, SB, MU, PN, FE. Funding acquisition: MU. All authors read and approved the final version of the manuscript.

## Data availability

Boxplots of the NPX values can be found open access on The Blood Resource of the Human Protein Atlas (https://www.proteinatlas.org/humanproteome/blood). Code is available on https://github.com/jjkenrick/pan-autoimmune-protein-profiling.git. Differential analysis and machine learning results available in supplementary material.

## Abbreviations

AUC: average area under the curve
HC: healthy controls
IDC: infectious disease controls
IIM: idiopathic inflammatory myopathies
PEA: proximity extension assay
RA: rheumatoid arthritis
ROC: receiver operating characteristic
SARDs: systemic autoimmune rheumatic diseases
SjD: Sjögren disease
SLE: systemic lupus erythematous
SSc: systemic sclerosis
NPX: normalized protein expression

## SUPPLEMENTARY INFORMATION

### SUPPLEMENTARY FIGURES

**Supplementary Fig. S1:**
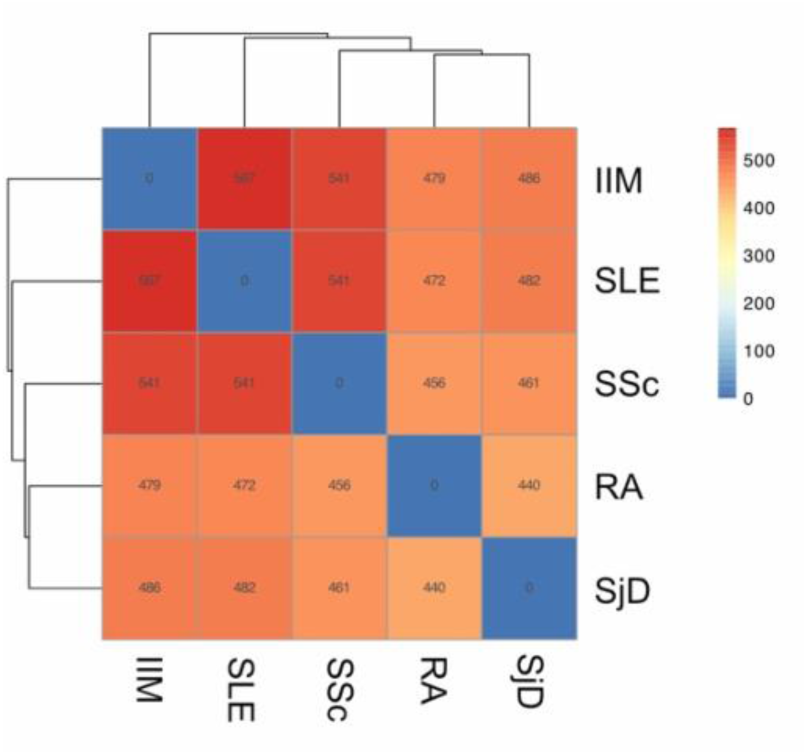
Higher protein levels across diseases compared to healthy controls. Heat map showing the number of overlapping proteins from each individual SARD that had higher levels (adj.P.val < 0.01 & logFC > 0.25) compared to the healthy control.

**Supplementary Fig. S2:**
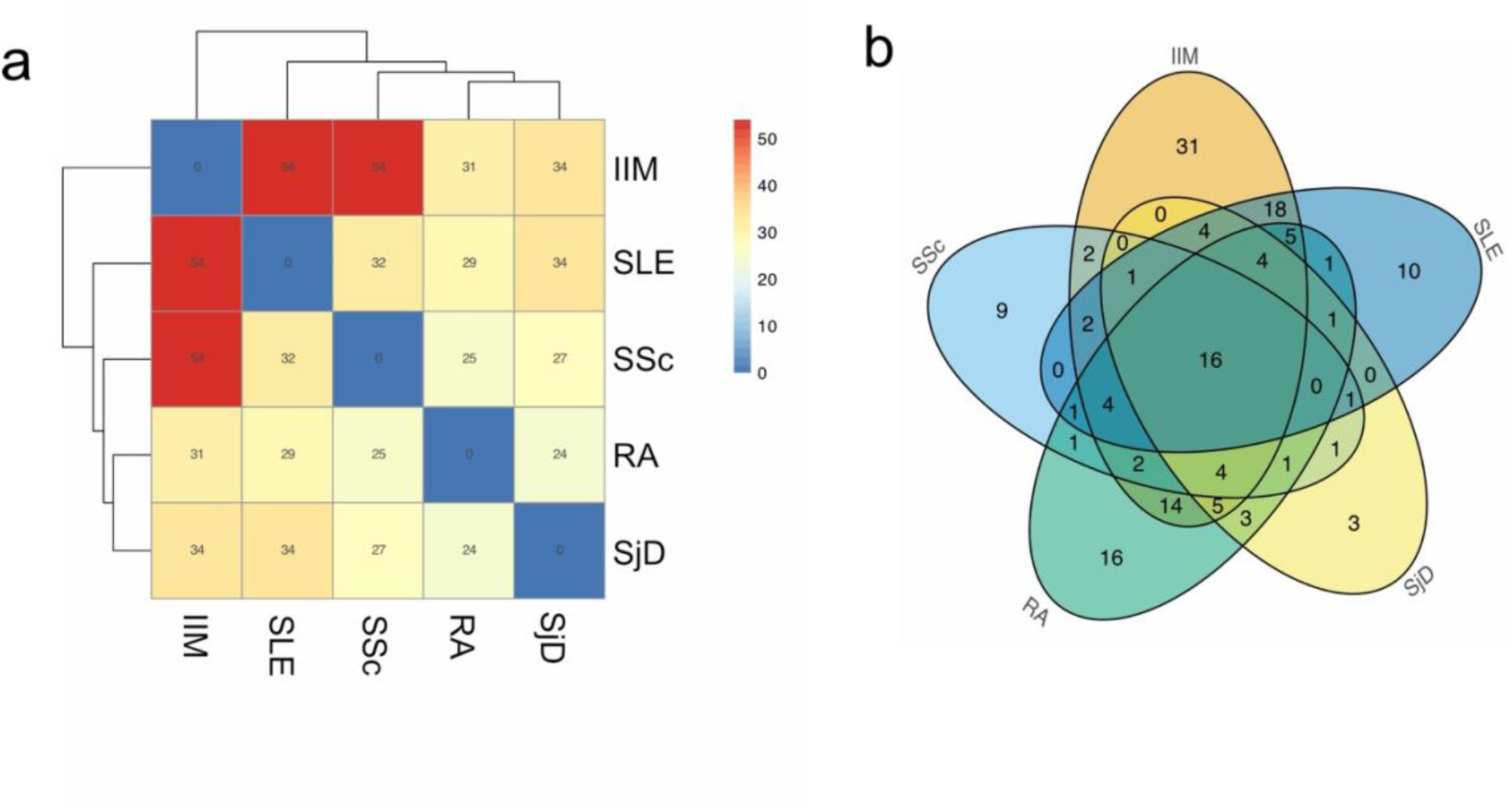
Lower proteins levels across diseases compared to healthy controls. **a**) Heat map showing the number of overlapping proteins from each individual SARD that had lower levels (adj.P.val < 0.01, logFC < –0.25) compared to the healthy control; **b**) Venn diagram showing the number of overlapping proteins from each individual SARD that had lower levels (adj.P.val < 0.01, logFC < –0.25) compared to the healthy control.

**Supplementary Fig. S3:**
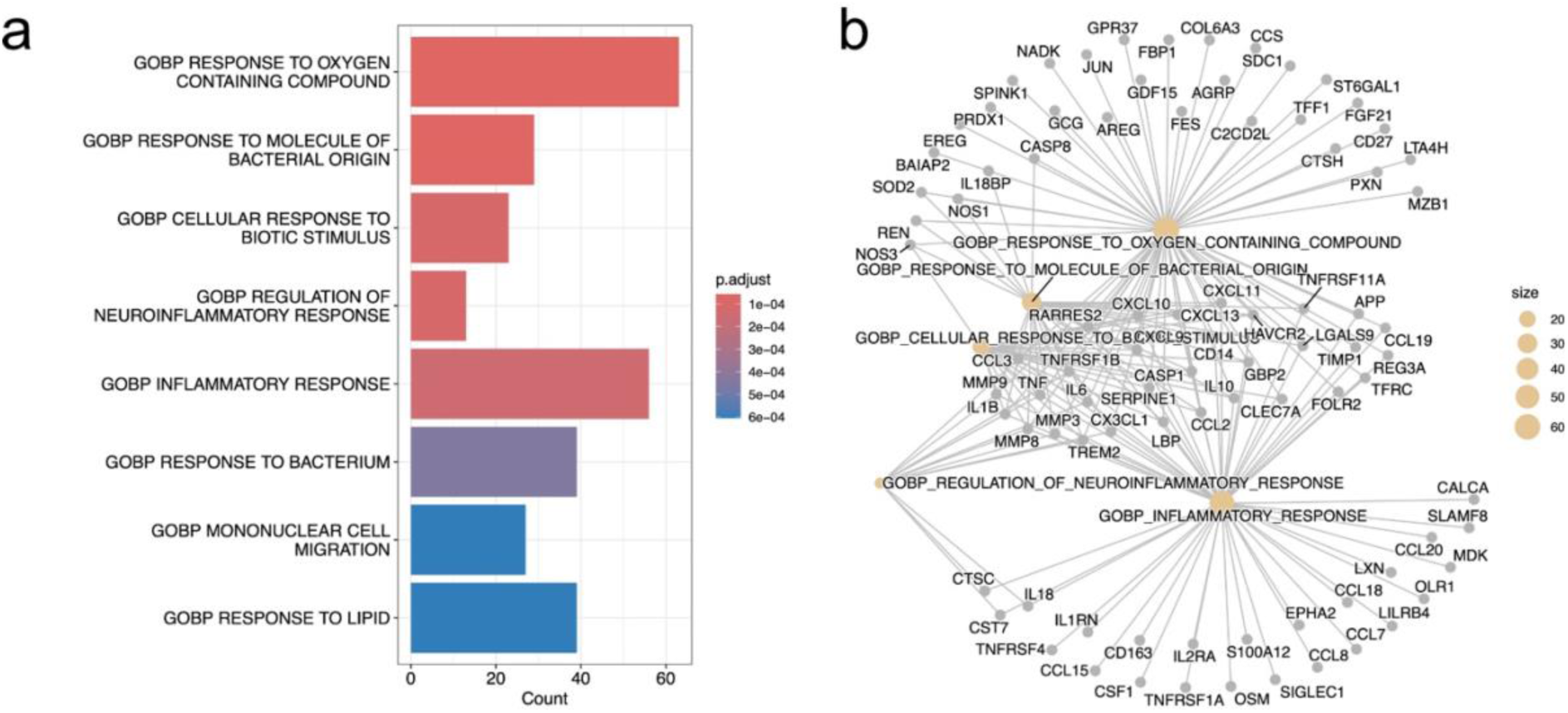
Grouped SARDs compared to healthy controls. Gene set enrichment of. **a**) barplot and **b**) gene-category network of proteins that had higher levels (adj.P.val < 0.01, logFC > 0.25) in the grouped SARDs against the healthy control.

**Supplementary Fig. S4:**
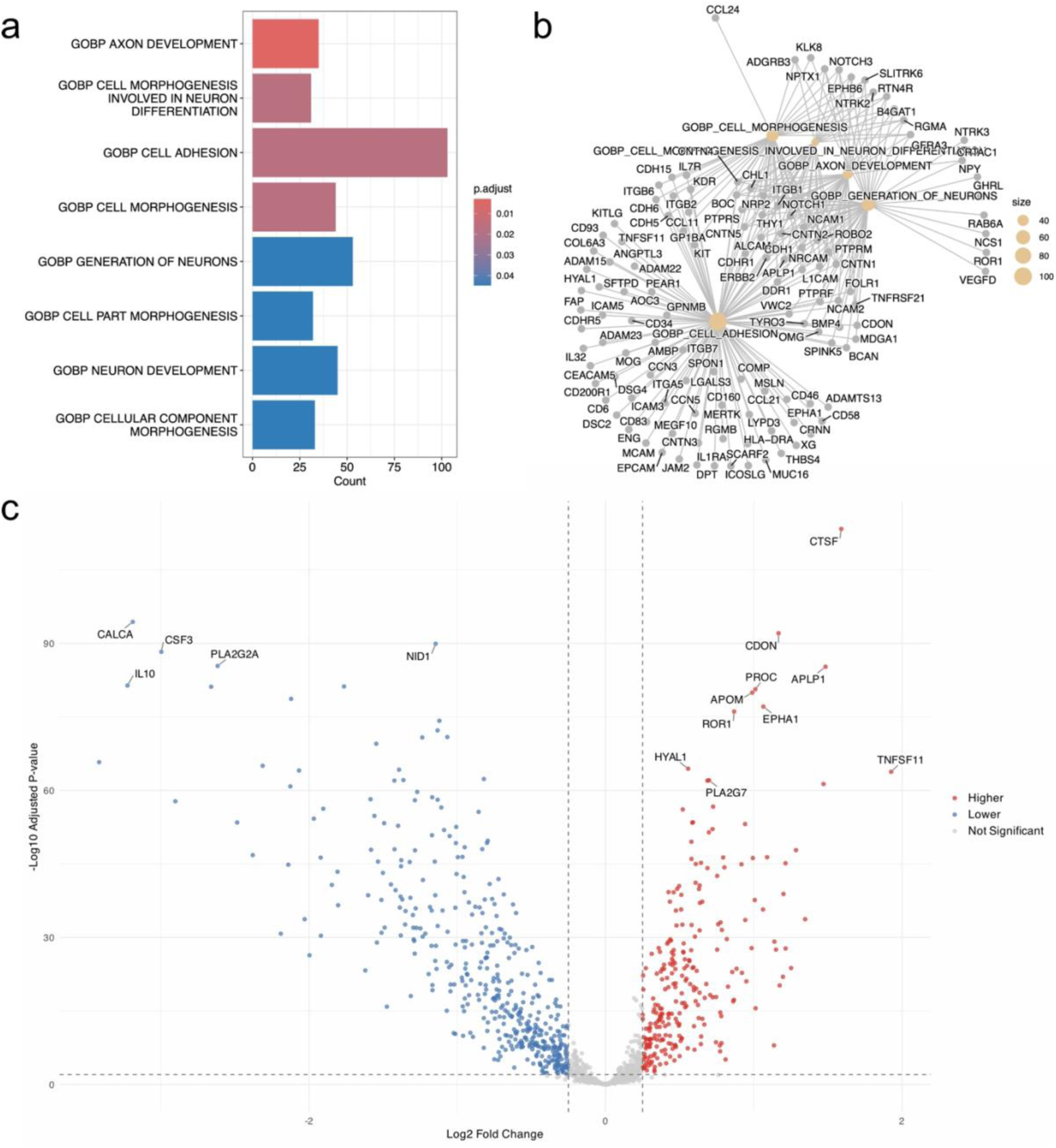
Grouped SARDs compared to infectious disease controls. Gene set enrichment barplot from proteins that had higher levels (adj.P.val < 0.01, logFC > 0.25) in the grouped SARDs than the infectious disease controls represented in **a**) barplot and **b**) gene-category network. **c**) Volcano plot highlighting proteins with differential levels in autoimmune cohort in comparison to grouped infectious disease. Proteins with significantly higher levels (adj.P.val < 0.01, logFC > 0.25) in autoimmune cohort are shown in red, and lower levels (adj.P.val < 0.01, logFC < –0.25) against infectious disease in blue.

**Supplementary Fig. S5:**
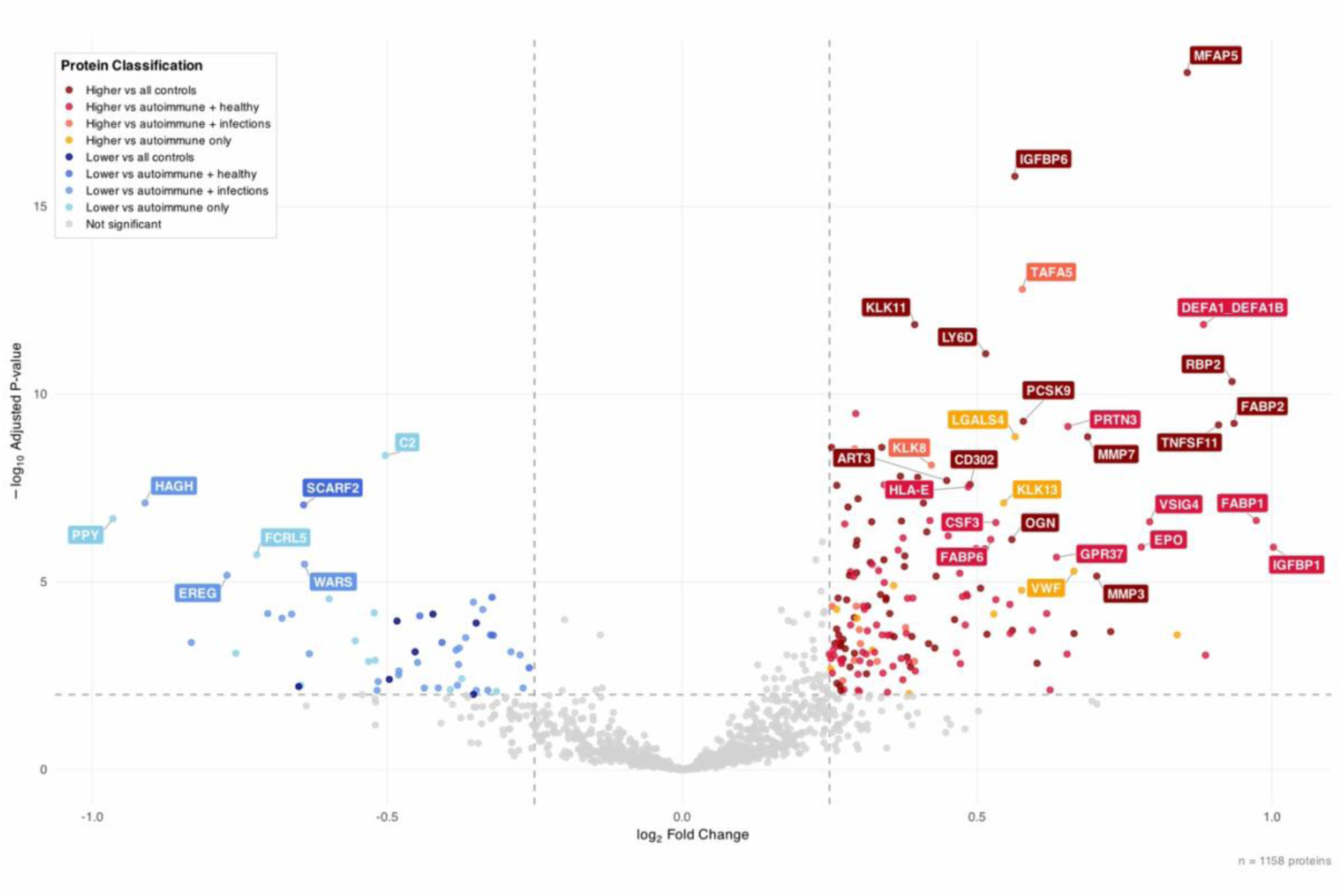
Volcano plot for comparison of systemic lupus erythematosus (SLE) to the grouped autoimmune diseases (adj.P.val < 0.01, |logFC| > 0.25), annotated by comparisons to healthy controls and grouped infectious disease controls.

**Supplementary Fig. S6:**
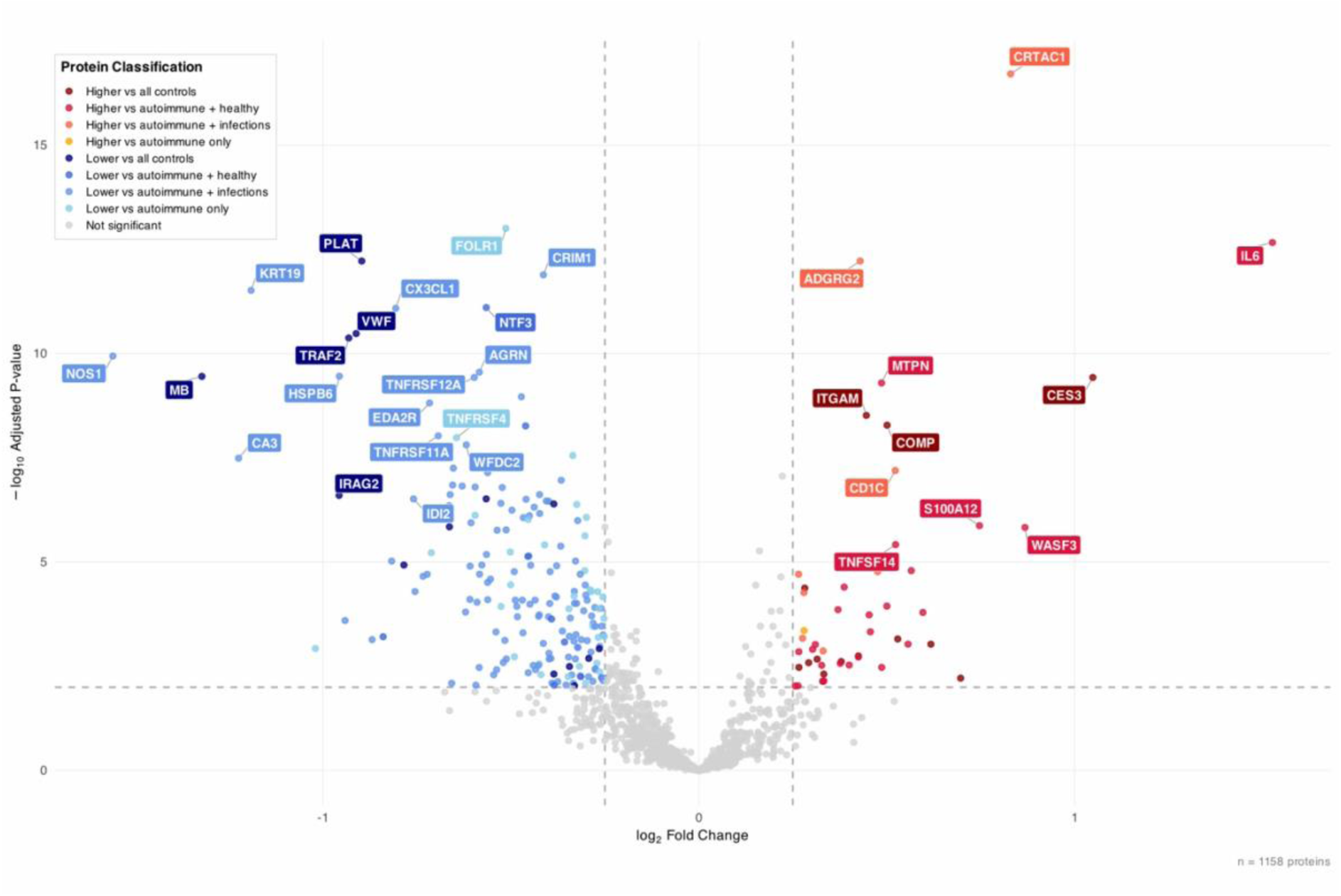
Volcano plot for comparison of rheumatoid arthritis (RA) to the grouped autoimmune diseases (adj.P.val<0.01, |logFC| > 0.25), annotated by comparisons to healthy controls and grouped infectious disease controls.

**Supplementary Fig. S7:**
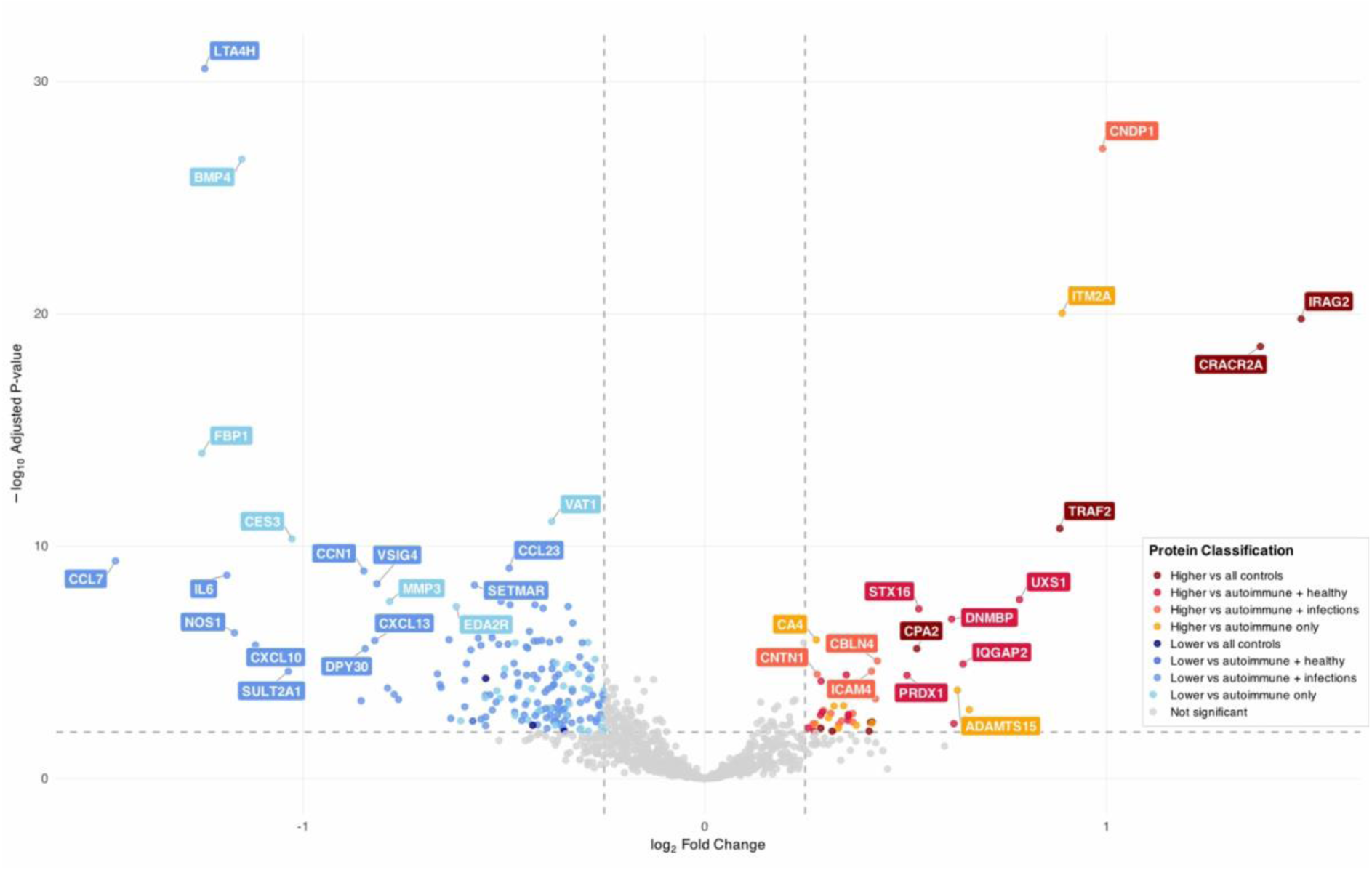
Volcano plot for comparison of Sjögren disease (SjD) to the grouped autoimmune diseases (adj.P.val<0.01, |logFC| > 0.25), annotated by comparisons to healthy control and grouped infectious disease controls.

**Supplementary Fig. S8:**
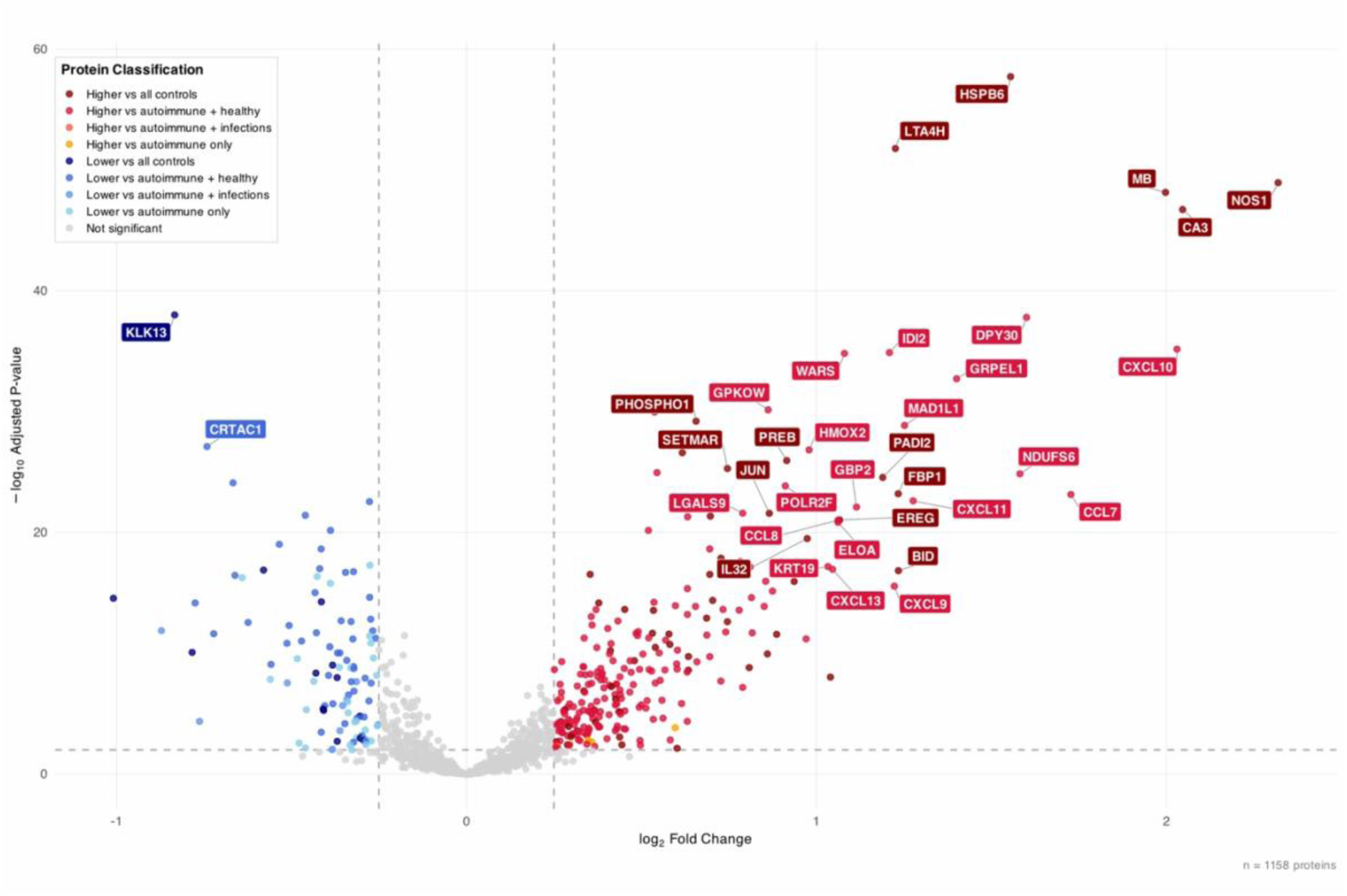
Volcano plot for comparison of idiopathic inflammatory myopathies (IIM) to the grouped autoimmune diseases (adj.P.val<0.01, |logFC| > 0.25), annotated by comparisons to healthy controls and grouped infectious disease controls.

